# Higher risk of mental health deterioration during the Covid-19 lockdown among students rather than non-students. The French Confins study

**DOI:** 10.1101/2020.11.04.20225706

**Authors:** Julie Arsandaux, Ilaria Montagni, Mélissa Macalli, Nathalie Texier, Mathilde Pouriel, Raphaël Germain, Adel Mebarki, Shérazade Kinouani, Marie Tournier, Stéphane Schück, Christophe Tzourio

## Abstract

**Background:** Covid-19 pandemic and its consequences have raised fears of its psychological impact. The objective of this study was to estimate the effect of student status on mental health conditions during Covid-19 general lockdown among adults in France.

**Methods:** Using cross-sectional data of the Confins cohort, we estimated the effect of student status on depressive and anxiety symptoms, suicidal thoughts and perceived stress using multivariate logistic regression analyses. Stratified models for college students and non-students were performed to identify associated population-specific factors.

**Results:** Among the 2260 included participants, students represented 59% (n=1335 vs 925 non-students) and 78% of the total sample were female. Student status was more frequently associated with depressive symptoms (adjusted OR(aOR)=1.58; 95%CI 1.17;2.14), anxiety symptoms (aOR=1.51; 95%CI 1.10;2.07), perceived stress (n=1919, aOR=1.70, 95%CI 1.26;2.29) and frequent suicidal thoughts (n=1919, aOR=1.57, 95%CI 0.97;2.53). Lockdown conditions that could be potentially aggravating on mental health like isolation had a higher impact on students than non-students.

**Limitations:** Participants were volunteers, which could limit generalisation of the findings. The cross-sectional design did not allow determining if lockdown impacted directly mental health or if there is another cause. However, we adjusted analyses with the history of psychiatric disorders, and factors related to lockdown conditions were associated with mental health disturbances.

**Conclusions:** College student’s mental health is of great importance in the context of the general lockdown set up during the pandemic. Follow-up and interventions should be implemented especially for those at high-risk (younger people and those with history of psychiatric disorders).

## INTRODUCTION

Young adults are particularly exposed to psychiatric disorders which often start in young adulthood. College students have been identified as a vulnerable population and have higher prevalence of psychiatric symptoms (e.g. depressive symptoms, anxiety symptoms and suicidal risk) than other adults (Kessler et al., 2007; Kovess-Masfety et al., 2016; Verger et al., 2010, 2009). In this population, and particularly in the first years at university, a high level of stress related to academic achievement, a low self-esteem, and depressive symptoms are frequently reported (Augesti et al., 2015; Beiter et al., 2015; Cheung et al., 2020).

The unprecedented context of 2020 with the major SARS-CoV-2 pandemic and its consequences have raised fears of its psychological impact in the population and more particularly in the fragile population of college students (Galea et al., 2020). In February 2020, the SARS-CoV-2 epidemic, causing the Covid-19 disease, hit Europe and France, one of the most affected countries in the world in number of cases and deaths (“WHO Coronavirus Disease (COVID-19) Dashboard | WHO Coronavirus Disease (COVID-19) Dashboard,” n.d.). To contain the spread of the epidemic, France established a general lockdown from March 17 to May 11, 2020. It has been shown in previous lockdown situations (e.g. SARS epidemic in 2013) that quarantine can have an impact on mental health (depression, post-traumatic stress symptoms, confusion, anger as well as suicide) by itself (Barbisch et al., 2015; Brooks et al., 2020).

Little information is available about the psychological impact of lockdown on college students and its risk of exacerbating their isolation and their psychological vulnerability (Beck et al., n.d.; Husky et al., 2020). A few studies have reported high prevalence of depressive symptoms, anxiety symptoms and stress during the Covid-19 lockdown among college students (Cao et al., 2020; Husky et al., 2020; Odriozola-González et al., 2020; Tang et al., 2020) but it is unkown whether the impact was different in this population compared to non-students adults. The objectives of this study were to estimate the effect of lockdown on mental health conditions (depressive symptoms, anxiety symptoms, suicidal thoughts and perceived stress) in college students and to compare their frequency and associated factors to a sample of non-students recruited in the same study.

## MATERIAL AND METHODS

### Data source, study design and study population

This study is based on the Confins e-cohort (www.confins.org), a prospective online population-based cohort study of adults in France set up since April 2020 during Covid-19 national lockdown and still ongoing. The objectives of the Confins cohort were: (1) to investigate the impact of the Covid-19 pandemic and the general lockdown, established in France between March and May 2020, on the well-being and mental health of the population; and (2) to explore opinions and beliefs of the population about the pandemic, its treatment and vaccines. A large communication campaign was deployed in France on social media and press, with a focus on specific populations (i.e. college students, health workers) reached also by emailing. The eligibility criteria were to be more than 18 and locked down in the French territory until the end of the general lockdown in France (May, 11, 2020). Enrolled participants signed in a secured web-site and complete questionnaires online. The baseline questionnaire collected socio-demographic information, medical history, lockdown conditions, mental health parameters, as well as opinions and beliefs. Thanks to the Confins cohort, direct comparison between students and non-students is possible (same recruitment and data collection for both populations). This study is based on baseline data collected during the general lockdown in France (until May, 11, 2020).

### Measures

#### Outcome: Mental health conditions

##### Depressive symptoms

Depressive symptoms were measured using the Patient Health Questionnaire-9 (PHQ-9) (Kroenke et al., 2001; Pfizer, n.d.) modified to assess symptoms within the last seven days (instead of the last 14 days) for close monitoring purpose. Items are rated from 0 to 3 according to increased frequency of experiencing difficulties in each area covered. Scores are summed and can range from 0 to 27. Higher scores represent higher depressive symptoms. We used the validated French version of the PHQ-9 (Arthurs et al., 2012). Since, distribution of the score was not normal, we used a validated cut-off of 10 to define the presence of depressive symptoms (Kroenke et al., 2001; Manea et al., 2012).

##### Anxiety symptoms

Anxiety symptoms were measured using the Generalized Anxiety Disorder- 7 (GAD-7) (Spitzer et al., 2006) modified to assessed symptoms within the last seven days (instead of the last 14 days). Items are rated from 0 to 3 according to increased frequency of experiencing difficulties in each area covered. Scores are summed and can range from 0 to 21. Higher scores represent higher anxiety symptoms. We used the validated French version of the GAD-7 (Micoulaud-Franchi et al., 2016). Since, distribution of the score was not normal, we used a validated cut-off of 10 to define the presence of anxiety symptoms (Kroenke et al., 2001).

##### Suicidal thoughts

Participants reported if they experienced suicidal thoughts during the last seven days (“*During the past 7 days, have you ever thought about killing yourself (suicidal ideas)*”: with the responses “*no, never*” and “*yes sometimes*” or “*yes, on multiple occasions*” considering together for analysis purpose).

##### Perceived stress

Participants rated their current stress on a 10 points-scale (“*How worried or stressed are you right now on a scale of 0-10*”), with 0 representing the lowest level of stress and 10 the highest. Since, distribution of the score was not normal, we used a cut-off of 7 to defined high perceived stress (corresponding approximately to the third quartile of the distribution).

##### Student status

Participants declared if they were currently college students or not and were asked about specific information according to their academic situation (e.g. cursus, university year) if they were students and their professional situation otherwise (having a high professional position and having a stable professional situation like time-undetermined work contract).

##### Covariates/other exposures

Sociodemographic information included sex and age (in years), familial situation (in a relationship or not) and education level collected differently for students and non-students and transformed for analysis into three categories: minor than a second-year university level or currently in their 1^st^ or 2^nd^ year for students; holding a second-year university level or currently in their 3^rd^ year for students; holding more than a second-year university level or currently in their 4^th^ or more year for students. Other variables possibly influencing on mental health were also recorded like working or studying in the medical field, having an history of psychiatric disorder (among depression, bipolar disorders, generalized anxiety), history of another disease at risk for severe forms of Covid-19 (among cardiovascular, respiratory, chronic digestive disease, cancer and diabetes). Other covariates/exposures were collected according to the student status or not. For students: having a paid activity (student job) and self-rated financial situation during childhood (correct, difficult or very difficult vs comfortable or very comfortable). For non-students: having a high professional position and having a stable professional situation (time-undetermined work contract).

Information related to the Covid-19 pandemic or lockdown were also recorded: week of inclusion (W14-15 corresponding to March 30 to April 12, W16-17 corresponding to April 13 to 26 and W18-19 corresponding to April 27 to May 11), being in lockdown in a high-risk region (defined by region that registered more than 10 death during the week 14), having acquaintance or family with Covid-19 or Covid-19 suspicion, being alone in lockdown accommodation, being in lockdown at home (in the same place than before lockdown), lockdown accommodation with an outdoor space (like a balcony or a garden), lockdown accommodation surface (in m^2^), having a pet, doing physical exercise during lockdown. Non-students declared if they were working remotely during lockdown or not.

### Statistical analysis

First, we described the study sample overall and in both student and non-student categories. Second, we estimated the effect of student status on each mental condition (i.e. depressive symptoms, anxiety symptoms, suicidal thoughts and perceived stress) using separated logistic regression models. Three multivariate models were built using a different adjustment: model 1 adjusted for age and sex, model 2 adjusted for age, sex and variables not related to the Covid- 19 pandemic or lockdown (i.e. being in a relationship, education level, working or studying for the medical sector, history of psychiatric disease, history of other disease at risk for Covid-19) and model 3 adjusted for age, sex, pre-cited variables not related to the Covid-19 pandemic or lockdown and variables related to the Covid-19 pandemic or lockdown (i.e. week of inclusion in the cohort, being in lockdown in a high-risk region, acquaintance or family with Covid-19 or Covid-19 suspicion, being in lockdown alone, lockdown accommodation with an outdoor space, having a pet, lockdown accommodation surface, physical activity during lockdown). We estimated odds ratio (OR), their 95% confidence interval (95% CI) and the p-value of the Wald test (and p-value of the type 3 test for categorical variables) related to the effect of student status. We performed secondary analysis restricting the sample to young adults (≤ 30 years old) to better take into account potential confusion by age.

Third, we computed a general model for each mental health conditions among students and non-students separately to investigate associated factors of these two populations. We performed the same model than primary analysis extending the list of exposure with all factors (model 3) and entering specific variables for students (i.e. paid activity and financial situation during childhood) and for non-students (i.e. high professional position, stable professional position, teleworking).

To test the robustness of the findings, we conducted sensitivity analyses using alternative cut- off for depressive, anxiety symptoms and perceived stress: (a) cut-off that represents severe depressive symptoms (PHQ-9 ≥ 15), severe anxiety symptoms (GAD-7 ≥ 15) and mild perceived stress (≥ 5), (b) cut-off that represents the full spectrum of depressive (minimal: 0-9; mild: 10-14; moderate: 15-19; severe: 20-27) and anxiety symptoms (minimal: 0-4; mild: 5-9; moderate: 10-14; severe: 15-21). We also performed sensitivity analyses among the complete case population to test the robustness of results regarding the imputation process.

Our missing data analysis procedures used missing at random assumptions. We used the multivariate imputation by chained equations (MICE) method of multiple multivariate imputation in SAS software (PROC MI and MIANALYZE) (Janssen et al., 2010; Rubin and Schenker, 1991; Schafer, 1997). We independently analysed 10 copies of the data, each with suitably imputed missing values, in the multivariate linear or logistic regression analyses. We averaged estimates of the variables to give a single mean estimate and adjusted standard errors according to Rubin’s rules. We imputed only data from covariates using covariate data and completed the imputation process with other data collected in the Confins cohort (having a child, housing type before and during the lockdown, self-rated health and self-rated quality of life before the lockdown). We performed additional imputations for the student subsample since paid activity and financial conditions during childhood had missing data.

We performed all analyses using the SAS statistical software (SAS V9.3).

## RESULTS

### Participants and sample description

Of the 2344 participants enrolled in the Confins cohort, 2309 were eligible for the study and 2260 were ultimately included in the primary analysis for depressive and anxiety symptoms and 1919 for suicidal thoughts and perceived stress (Fig1). Table 1 and Table S1 provided in supplementary material describe the study sample. Students represented 59% of the total sample (n=1335 vs 925 non-students). Mean age in the student sample was about 16 years lower (mean=23.3 vs 40.1) whereas sex ratio was similar (3/4 female) in both samples. Students had less frequently a partner (48.1% vs 76.3%) and were more frequently at risk for severe forms for Covid-19 (35.4% vs 24.0%). However, both populations were similar regarding history of psychiatric disorders (about 23%), education level (with a majority of more than a second-year university level) and proportion of individual working or studying in health domains (about 40%). Although non-students spent the lockdown more frequently in their usual place than students (88.2% vs 63.4%), the quality of lockdown accommodation (e.g. surface, outdoor space) was similar for both. Students were fewer than non-students to be in a high-risk region (11.3% vs 31.2%) and they were enrolled in the cohort later (mostly during weeks 16-17).

**Table 1.** Characteristics of the study population, Confins cohort, France, 2020

|  | **Total** |  | **Students** |  | **Non-students** |  |
| --- | --- | --- | --- | --- | --- | --- |
|  | **N=2260** |  | **N=1335** |  | **N=925** |  |
| Female, n (%) | 1759 | (77.8) | 1065 | (79.8) | 694 | (75.0) |
| Age, mean (STD) | 30.2 | (12.8) | 23.3 | (3.9) | 40.1 | (14.5) |
| <30 years old, n (%) | 1600 | (70.8) | 1281 | (96.0) | 319 | (34.5) |
| In a relationship, n (%) | 1348 | (59.7) | 642 | (48.1) | 706 | (76.3) |
| Education level, n (%) (N=2259) | 2259 |  | 1335 |  | 924 |  |
| < Second-year university level | 462 | (20.5) | 251 | (18.8) | 211 | (22.8) |
| Second-year university level | 393 | (17.4) | 216 | (16.2) | 177 | (19.2) |
| > Second-year university level | 1404 | (62.2) | 868 | (65.0) | 536 | (58.0) |
| Working or studying for the medical sector, n (%) | 882 | (39.0) | 507 | (38.0) | 375 | (40.5) |
| History of psychiatric disease, n (%)<br>(missing data: 209/186) | 425 | (22.8) | 254 | (22.1) | 171 | (23.9) |
| History of other disease at risk for covid-19*, n (%) (missing data: 197/144) | 544 | (28.4) | 286 | (24.0) | 258 | (35.4) |
| Weeks of inclusion in the cohort, n (%)<br>(N=2259) |  |  |  |  |  |  |
| W14-15 | 676 | (29.9) | 264 | (19.8) | 412 | (45.6) |
| W16-17 | 1306 | (57.8) | 862 | (64.6) | 444 | (48.1) |
| W18-19 | 277 | (12.3) | 209 | (15.7) | 68 | (7.4) |
| Being in lockdown in a high-risk region, n (%) (N=2254) | 440 | (19.5) | 151 | (11.3) | 289 | (31.2) |
| Lockdown accommodation with an outdoor space, n (%) | 1796 | (79.5) | 1039 | (77.8) | 757 | (81.8) |
| Having a pet, n (%) | 1002 | (44.3) | 616 | (46.1) | 386 | (41.7) |
| Lockdown accommodation surface (in m<sup>2</sup>),<br>mean (STD) (missing data: 0/8) | 121.2 | (239.0) | 124.5 | (255.8) | 116.4 | (212.6) |
| Acquaintance or family with Covid-19 or<br>Covid-19 suspicion, n (%) | 963 | (42.6) | 556 | (41.7) | 407 | (44.0) |
| Being in lockdown alone, n (%) | 368 | (16.3) | 221 | (16.6) | 147 | (15.9) |
| Physical exercise during lockdown, n (%) | 1863 | (82.4) | 1081 | (81.0) | 782 | (84.5) |
| Being in lockdown at home, n (%) | 1662 | (73.5) | 846 | (63.4) | 816 | (88.2) |

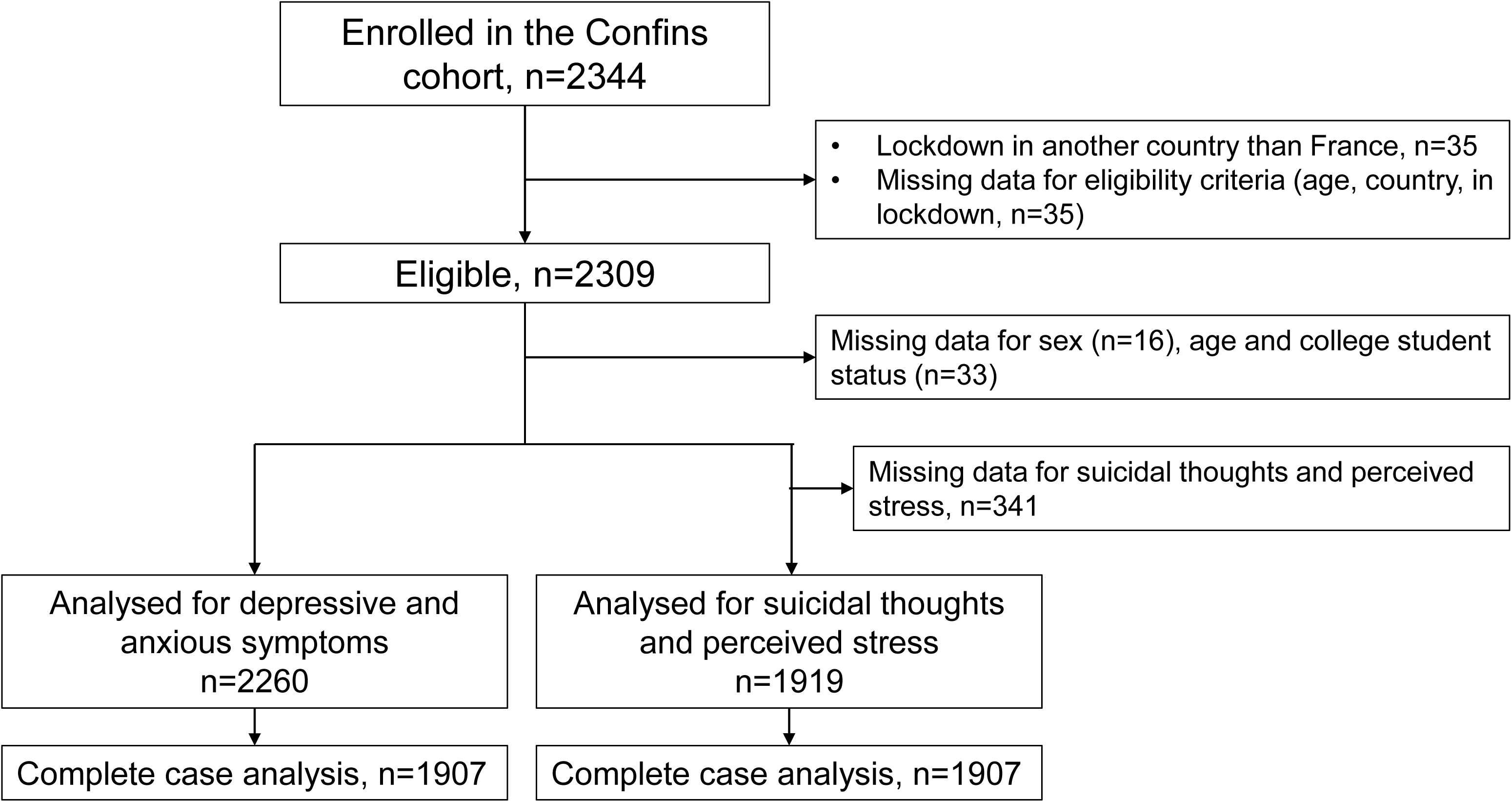

### Student status and mental health conditions

During lockdown period, mental health conditions were different between the two populations. Students presented more frequently depressive symptoms (32.5% vs 16.2%), anxiety symptoms (24.0% vs 14.7%), suicidal thoughts (11.7% vs 7.6%) and perceived stress (33.1% vs 22.1%) than non-students (Table 2). In multivariate models, student status was associated with an increased probability to report depressive symptoms (OR fully adjusted=1.58; 95%CI=1.17;2.14), anxiety symptoms (OR fully adjusted=1.51; 95%CI=1.10;2.07), and perceived stress (OR fully adjusted=1.70, 95%CI=1.26;2.29), independently from covariates related or not to the Covid-19 pandemic or lockdown (Table 3). For suicidal thoughts, the odd-ratios were in the same range (OR fully adjusted=1.57; 95%CI=0.97;2.53) but did not reach significance.

**Table 2.** Mental health conditions during lockdown, Confins cohort, France, 2020

|  | Total |  | Students |  | Non-students |  |
| --- | --- | --- | --- | --- | --- | --- |
|  | N=2260 |  | N=1335 |  | N=925 |  |
| Depressive symptoms |  |  |  |  |  |  |
| PHQ-9 score ≥ 10, n (%) | 584 | (25.8) | 434 | (32.5) | 150 | (16.2) |
| Anxiety symptoms |  |  |  |  |  |  |
| GAD-7 score ≥ 10, n (%) | 457 | (20.2) | 321 | (24.0) | 136 | (14.7) |
| Suicidal thoughts, n (%) (missing data: 195/142) | 194 | (10.1) | 139 | (11.7) | 55 | (7.6) |
| Perceived stress (missing data: 195/142) |  |  |  |  |  |  |
| Perceived stress ≥ 7, n (%) | 555 | (28.9) | 394 | (33.1) | 161 | (22.1) |

**Table 3.**
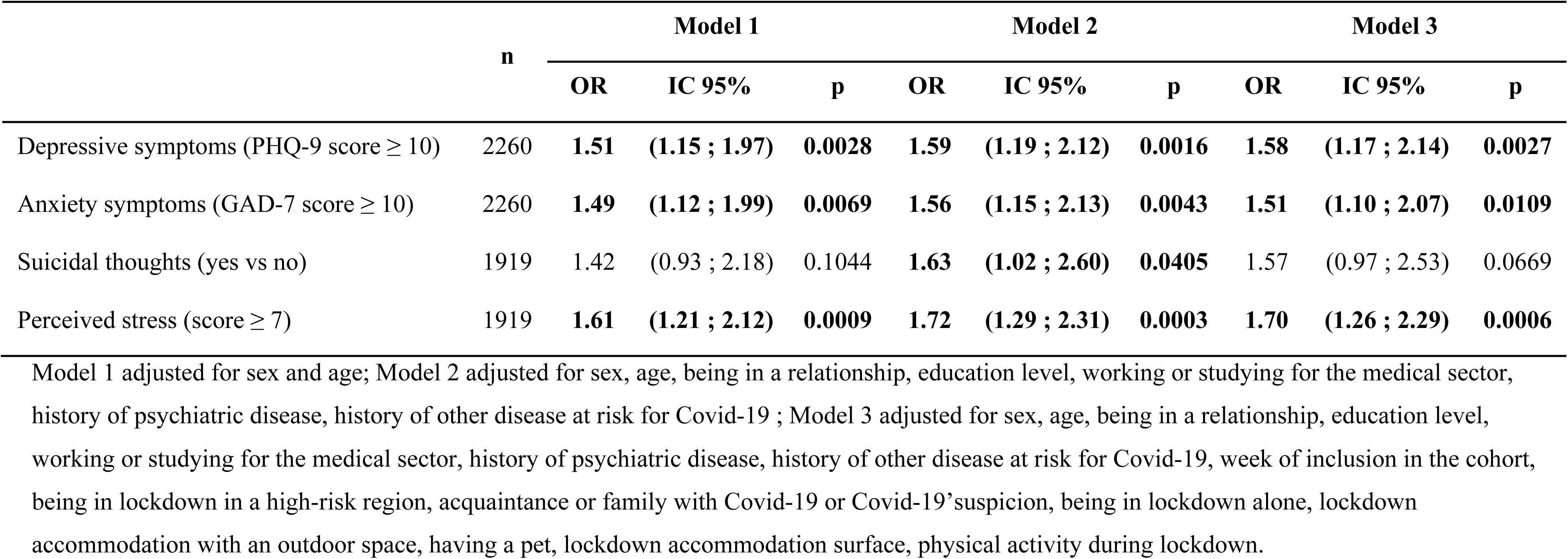
Effect of college student status on mental health conditions during Covid-19 lockdown, estimated with multivariate logistic regression analysis with imputed data on covariates, Confins cohort, France, 2020.

Sensitivity analyses showed consistent results when analyses were performed among the complete case population and with different cut-offs (results presented in Table S2 in supplementary material). When both samples were restricted to young adults (≤ 30), students still had a higher risk of depressive symptoms (OR fully adjusted=1.54; 95%CI=1.03;2.29), anxiety symptoms (OR fully adjusted=1.60; 95%CI=1.04;2.45) and perceived stress (OR fully adjusted=1.59; 95%CI=1.07;2.34) (results presented in Table S3 supplementary material).

### Stratified analyses on influencing factors

Students reported consistently more frequently depressive symptoms whatever the strata: female, male, with a history of psychiatric disorders, or according to various lockdown conditions (e.g. being in lockdown alone) (Fig2). In explicative models for mental health conditions, we found shared factors between students and non-students: past history of psychiatric disorders (e.g. for depressive symptoms: OR=2.80, 95%CI=2.06;3.82 among students and OR=3.10, 95%CI=1.99;4.81 among non-students) and physical exercise during lockdown which was associated with less frequent mental health disturbances (e.g. fo depressive symptoms: OR=0.61, 95%CI=0.44;0.83 among students and OR=0.52, 95%CI=0.32;0.83 among non-students). Certain aggravating factors of lockdown were associated with more frequent mental health disturbances, especially among students. Indeed, being isolated in lockdown was associated with depressive symptoms only among students (OR=1.54, 95%CI 1.04;2.28 among students vs OR=0.90, 95%CI 0.50;1.63 among non-students). In contrast, other factors were protector, like being in lockdown at home only for students (OR=0.75, 95%CI 0.57;0.99 among students vs OR=1.88, 95%CI 0.85;4.18 among non-students). Detailed results of explicative models are available in Table S4 and S5 in supplementary material.

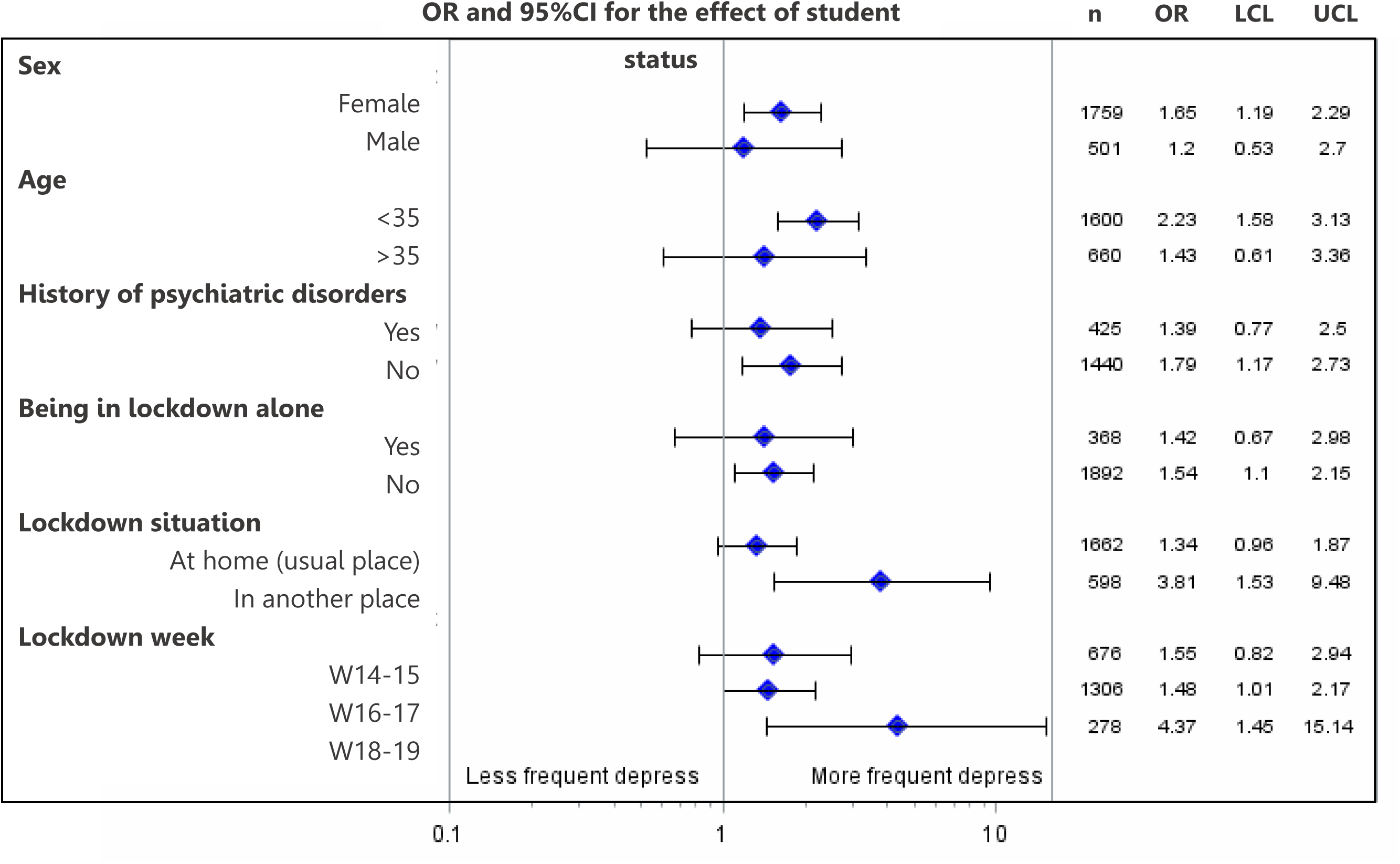

## DISCUSSION

In this large sample of adults observed during the general lockdown, high level of mental health disturbances was observed. Students had a much higher prevalence of mental health problems (more depressive symptoms, anxiety symptoms and perceived stress) than non-students. In multivariate models adjusting for a large variety of potential confounding factors, students had a more than 50% increased risk of mental health problems compared to non-students. This increased risk was also observed in participants with or without a past history of psychiatric diseases, in the various strata of lockdown situations, or when both samples were restricted to young adults. Finally, lockdown conditions that could be potentially aggravating on mental health like isolation had a higher impact on students.

### Interpretations

High frequency of mental health disturbances found in our study corroborates previous research conducted separately among students and other adults. Among the general non-student population, several studies reported a prevalence between 17% and 28% for depressive symptoms, between 14% and 45% for anxiety symptoms and between 8% and 27% for stress (Bäuerle et al., n.d.; Choi et al., 2020; González-Sanguino et al., 2020; Gualano et al., 2020; Mazza et al., 2020; Ozamiz-Etxebarria et al., 2020; Wang et al., 2020). Among college students, several studies reported a prevalence during the lockdown ranging between 9% and 21% for depressive symptoms, between 25% and 34% for anxiety symptoms and 28% for stress (Cao et al., 2020; Odriozola-González et al., 2020; Tang et al., 2020). Our study directly compared students and non-students in the same cohort and brings strong evidence for a higher risk of mental health disturbances among students rather than non-students during general lockdown.

Mental health during lockdown was associated with lockdown conditions, especially among students. Indeed, estimated frequency of mental health disturbances are higher during the general lockdown (in our study and in above-mentioned studies) compared to previous research conducted before Covid-19 pandemic and lockdown among students (between 11% and 20% of depressive symptoms, 15% of anxiety symptoms, and 11% of stress) (Beiter et al., 2015; Ibrahim et al., 2013; Rotenstein et al., 2016). Furthermore, lockdown conditions were associated with mental health among students in our study, showing that lockdown represents a high-risk period for students. Lockdown conditions were yet partially explored in the literature with an association found between mental health and having relative or acquaintance who got Covid-19 (Cao et al., 2020; Tang et al., 2020).

To explain this higher mental health risk for individuals during general lockdown, several mechanisms could be discussed: disturbance in mood homeostasis during lockdown (i.e. failure to stabilize mood via mood-modifying activities) (Taquet et al., 2020), lack of social and familial support (Gariépy et al., 2016; Kmietowicz, 2020; Pössel et al., 2018) or specific vulnerability of young adults and students already explored in the literature that could be exacerbated by pandemic and lockdown (e.g. worries about the future and employment) (Hunt and Eisenberg, 2010; Tang et al., 2020; Twenge et al., 2017).

### Strengths and limitations

The strengths of our study include the large sample, the standardised assessment tools used for mental health conditions and broad adjustment for other factors (related or not the Covid-19 pandemic or lockdown). This study profits recruitment of these two populations in the same cohort as well as similar data collection that allow direct comparison. Some limitations should however be taken into consideration. First, a sampling bias could have arisen since participants were volunteers, which could limit generalisation of the findings. Regarding mental health disturbances, the consequences appear however limited as frequencies were consistent with existing literature. Second, the cross-sectional design did not allow determining if lockdown impacted directly mental health or if there is another cause. However, we considered the history of psychiatric disorders that is an important confounder, and factors related to lockdown conditions were associated with mental health disturbances, suggesting that lockdown should have an impact on mental health especially for students. Longitudinal studies have been set up in the general population and have showed a deterioration of mental health during lockdown (Pierce et al., 2020). Similar studies are needed among students to establish clearly if lockdown can impact mental health in the short and in the long term.

### Implications

From a public health perspective, this study confirms that students were a vulnerable population for mental health disturbances during pandemic period and lockdown, suggesting that screening might be useful to offer adapted support. Besides, for preventive purpose in case of a new lockdown perspective, support and interventions adapted to students should be created and evaluated. Especially psychological support could be provided by University health student services both online and through crisis hotlines. Peers could also organize self-help groups and detect students with particular distress.

From a research perspective, we recommend to explore two priority areas. First, research is warranted in order to clarify causality of the relationship between lockdown and mental health disturbances among students. To this end, longitudinal mental health data covering a period before, during and after lockdown would be necessary. Second, it appears relevant to better understand the mechanisms underlying the specific vulnerability of students, i.e. identifying intermediate factors and especially those that can be modified. Besides, qualitative studies or mixed design (i.e. mix of quantitative and qualitative data) may be useful to better understand both psychologic disorders students experienced during this period and solutions some of them put in place to better cope with depression, anxiety, stress and suicidal thoughts.

## Data Availability

All data generated or analysed during this study are included in this published article. The full dataset is available upon request from the Confins cohort team (www.confins.org).

## ETHICS APPROVAL

The study follows the principles of the Declaration of Helsinki and the collection, storage and analysis of the data comply with the General Data Protection Regulation (EU GDPR).

The study was approved by the French Committee for the Protection of Individuals (Comité de Protection des Personnes – CPP IDF X, nr. 46-2020) and by the National Commission on Informatics and Liberty (Commission Nationale de l’Informatique et des Libertés - CNIL, nr. MLD/MFI/AR205600).

### Consent to participate

Students were informed on the nature and purpose of the study and provided on-line consent.

## AUTHOR APROUVAL

All authors have seen and approved the manuscript.

## AUTHORS’ CONTRIBUTIONS

C. Tzourio and S. Schück are the principal investigators of the Confins cohort. J. Arsandaux developed the study concept and the study design. I. Montagni, M. Macalli, S. Kinouani and M. Tournier participated actively in the creation of the Confins questionnaires and brought epidemiological, psychological and psychiatric expertise to the interpretation of the results. N. Texier, M. Pouriel, R. Germain and A. Mebarki brought technical expertise for the electronic case report form, data-management and statistical considerations. J. Arsandaux performed the data analysis and drafted the manuscript. All co-authors provided critical revisions and approved the final version of the manuscript for submission.

## ACKNOWLEDGEMENTS

The authors are indebted to the participants of the Confins cohort for their commitment and cooperation and to the i-Share operational team (Elena Milesi, Marie Mougin, Edwige Pereira, Garance Perret, Clothilde Pollet, Aude Pouymayou,), Kappa Santé (Christophe Brunat, David Clément, Clément Leclercq) and Kap Code (Vanessa Marie-Joseph, Juliette Olivier, Salma Rzin) for their expert contribution and assistance, especially in this particular context of pandemic and lockdown.

## REFERENCES

Arthurs, E., Steele, R.J., Hudson, M., Baron, M., Thombs, B.D., (CSRG) Canadian Scleroderma Research Group, 2012. Are Scores on English and French Versions of the PHQ-9 Comparable? An Assessment of Differential Item Functioning. PLoS ONE 7, e52028. https://doi.org/10.1371/journal.pone.0052028

Augesti, G., Lisiswanti, R., Saputra, O., Nisa, K., 2015. DIFFERENCES IN STRESS LEVEL BETWEEN FIRST YEAR AND LAST YEAR MEDICAL STUDENTS IN MEDICAL FACULTY OF LAMPUNG UNIVERSITY. J. Major. 4.

Barbisch, D., Koenig, K.L., Shih, F.Y., 2015. Is There a Case for Quarantine? Perspectives from SARS to Ebola. Disaster Med. Public Health Prep. https://doi.org/10.1017/dmp.2015.38

Bäuerle, A., Teufel, M., Musche, V., Weismüller, B., Kohler, H., Hetkamp, M., Dörrie, N., Schweda, A., Skoda, E.-M., n.d. Increased generalized anxiety, depression and distress during the COVID-19 pandemic: a cross-sectional study in Germany. J. Public Health. https://doi.org/10.1093/pubmed/fdaa106

Beck, F., Léger, D., Fressard, L., Peretti‐Watel, P., Verger, P., n.d. Covid-19 health crisis and lockdown associated with high level of sleep complaints and hypnotic uptake at the population level. J. Sleep Res. n/a, e13119. https://doi.org/10.1111/jsr.13119

Beiter, R., Nash, R., McCrady, M., Rhoades, D., Linscomb, M., Clarahan, M., Sammut, S., 2015. The prevalence and correlates of depression, anxiety, and stress in a sample of college students. J. Affect. Disord. 173, 90–96. https://doi.org/10.1016/j.jad.2014.10.054

Brooks, S.K., Webster, R.K., Smith, L.E., Woodland, L., Wessely, S., Greenberg, N., Rubin, G.J., 2020. The psychological impact of quarantine and how to reduce it: rapid review of the evidence. The Lancet. https://doi.org/10.1016/S0140-6736(20)30460-8

Cao, W., Fang, Z., Hou, G., Han, M., Xu, X., Dong, J., Zheng, J., 2020. The psychological impact of the COVID-19 epidemic on college students in China. Psychiatry Res. 287, 112934. https://doi.org/10.1016/j.psychres.2020.112934

Cheung, Dr.K., Tam, Dr.K.Y., Tsang, Ms.H., Zhang, Dr.L.W., Lit, Dr.S.W., 2020. Depression, anxiety and stress in different subgroups of first-year university students from 4-year cohort data. J. Affect. Disord. 274, 305–314. https://doi.org/10.1016/j.jad.2020.05.041

Choi, E.P.H., Hui, B.P.H., Wan, E.Y.F., 2020. Depression and Anxiety in Hong Kong during COVID-19. Int. J. Environ. Res. Public. Health 17, 3740. https://doi.org/10.3390/ijerph17103740

Galea, S., Merchant, R.M., Lurie, N., 2020. The Mental Health Consequences of COVID-19 and Physical Distancing: The Need for Prevention and Early Intervention. JAMA Intern. Med. 180, 817–818. https://doi.org/10.1001/jamainternmed.2020.1562

Gariépy, G., Honkaniemi, H., Quesnel-Vallée, A., 2016. Social support and protection from depression: systematic review of current findings in Western countries. Br. J. Psychiatry 209, 284–293. https://doi.org/10.1192/bjp.bp.115.169094

González-Sanguino, C., Ausín, B., Castellanos, M.Á., Saiz, J., López-Gómez, A., Ugidos, C., Muñoz, M., 2020. Mental health consequences during the initial stage of the 2020 Coronavirus pandemic (COVID-19) in Spain. Brain. Behav. Immun. 87, 172–176. https://doi.org/10.1016/j.bbi.2020.05.040

Gualano, M.R., Lo Moro, G., Voglino, G., Bert, F., Siliquini, R., 2020. Effects of Covid-19 Lockdown on Mental Health and Sleep Disturbances in Italy. Int. J. Environ. Res. Public. Health 17, 4779. https://doi.org/10.3390/ijerph17134779

Hunt, J., Eisenberg, D., 2010. Mental Health Problems and Help-Seeking Behavior Among College Students. J. Adolesc. Health 46, 3–10. https://doi.org/10.1016/j.jadohealth.2009.08.008

Husky, M.M., Kovess-Masfety, V., Swendsen, J.D., 2020. Stress and anxiety among university students in France during Covid-19 mandatory confinement. Compr. Psychiatry 102, 152191. https://doi.org/10.1016/j.comppsych.2020.152191

Ibrahim, A.K., Kelly, S.J., Adams, C.E., Glazebrook, C., 2013. A systematic review of studies of depression prevalence in university students. J. Psychiatr. Res. 47, 391–400. https://doi.org/10.1016/j.jpsychires.2012.11.015

Janssen, K.J.M., Donders, A.R.T., Harrell, F.E., Vergouwe, Y., Chen, Q., Grobbee, D.E., Moons, K.G.M., 2010. Missing covariate data in medical research: to impute is better than to ignore. J. Clin. Epidemiol. 63, 721–727. https://doi.org/10.1016/j.jclinepi.2009.12.008

Kessler, R.C., Amminger, G.P., Aguilar-Gaxiola, S., Alonso, J., Lee, S., Üstün, T.B., 2007. Age of onset of mental disorders: A review of recent literature. Curr. Opin. Psychiatry. https://doi.org/10.1097/YCO.0b013e32816ebc8c

Kmietowicz, Z., 2020. Rules on isolation rooms for suspected covid-19 cases in GP surgeries to be relaxed. BMJ. https://doi.org/10.1136/bmj.m707

Kovess-Masfety, V., Leray, E., Denis, L., Husky, M., Pitrou, I., Bodeau-Livinec, F., 2016. Mental health of college students and their non-college-attending peers: Results from a large French cross-sectional survey. BMC Psychol. https://doi.org/10.1186/s40359-016-0124-5

Kroenke, K., Spitzer, R.L., Williams, J.B.W., 2001. The PHQ-9. J. Gen. Intern. Med. 16, 606–613. https://doi.org/10.1046/j.1525-1497.2001.016009606.x

Manea, L., Gilbody, S., McMillan, D., 2012. Optimal cut-off score for diagnosing depression with the Patient Health Questionnaire (PHQ-9): a meta-analysis. CMAJ Can. Med. Assoc. J. J. Assoc. Medicale Can. 184, E191–196. https://doi.org/10.1503/cmaj.110829

Mazza, C., Ricci, E., Biondi, S., Colasanti, M., Ferracuti, S., Napoli, C., Roma, P., 2020. A Nationwide Survey of Psychological Distress among Italian People during the COVID-19 Pandemic: Immediate Psychological Responses and Associated Factors. Int. J. Environ. Res. Public. Health 17, 3165. https://doi.org/10.3390/ijerph17093165

Micoulaud-Franchi, J.-A., Lagarde, S., Barkate, G., Dufournet, B., Besancon, C., Trébuchon-Da Fonseca, A., Gavaret, M., Bartolomei, F., Bonini, F., McGonigal, A., 2016. Rapid detection of generalized anxiety disorder and major depression in epilepsy: Validation of the GAD-7 as a complementary tool to the NDDI-E in a French sample. Epilepsy Behav. 57, 211–216. https://doi.org/10.1016/j.yebeh.2016.02.015

Odriozola-González, P., Planchuelo-Gómez, Á., Irurtia, M.J., de Luis-García, R., 2020. Psychological effects of the COVID-19 outbreak and lockdown among students and workers of a Spanish university. Psychiatry Res. 290, 113108. https://doi.org/10.1016/j.psychres.2020.113108

Ozamiz-Etxebarria, N., Idoiaga Mondragon, N., Dosil Santamaría, M., Picaza Gorrotxategi, M., 2020. Psychological Symptoms During the Two Stages of Lockdown in Response to the COVID-19 Outbreak: An Investigation in a Sample of Citizens in Northern Spain. Front. Psychol. 11. https://doi.org/10.3389/fpsyg.2020.01491

Pfizer, n.d. Patient health questionnaire (PHQ) screeners [WWW Document]. URL http://www.phqscreeners.com (xaccessed 6.4.19).

Pierce, M., Hope, H., Ford, T., Hatch, S., Hotopf, M., John, A., Kontopantelis, E., Webb, R., Wessely, S., McManus, S., Abel, K.M., 2020. Mental health before and during the COVID-19 pandemic: a longitudinal probability sample survey of the UK population. Lancet Psychiatry 0. https://doi.org/10.1016/S2215-0366(20)30308-4

Pössel, P., Burton, S.M., Cauley, B., Sawyer, M.G., Spence, S.H., Sheffield, J., 2018. Associations between Social Support from Family, Friends, and Teachers and depressive Symptoms in Adolescents. J. Youth Adolesc. 47, 398–412. https://doi.org/10.1007/s10964-017-0712-6

Rotenstein, L.S., Ramos, M.A., Torre, M., Segal, J.B., Peluso, M.J., Guille, C., Sen, S., Mata, D.A., 2016. Prevalence of Depression, Depressive Symptoms, and Suicidal Ideation Among Medical Students: A Systematic Review and Meta-Analysis. JAMA 316, 2214–2236. https://doi.org/10.1001/jama.2016.17324

Rubin, D.B., Schenker, N., 1991. Multiple imputation in health-are databases: An overview and some applications. Stat. Med. 10, 585–598. https://doi.org/10.1002/sim.4780100410

Schafer, J.L., 1997. Analysis of Incomplete Multivariate Data. CRC Press.

Spitzer, R.L., Kroenke, K., Williams, J.B.W., Löwe, B., 2006. A Brief Measure for Assessing Generalized Anxiety Disorder: The GAD-7. Arch. Intern. Med. 166, 1092–1097. https://doi.org/10.1001/archinte.166.10.1092

Tang, W., Hu, T., Hu, B., Jin, C., Wang, G., Xie, C., Chen, S., Xu, J., 2020. Prevalence and correlates of PTSD and depressive symptoms one month after the outbreak of the COVID-19 epidemic in a sample of home-quarantined Chinese university students. J. Affect. Disord. 274, 1–7. https://doi.org/10.1016/j.jad.2020.05.009

Taquet, M., Quoidbach, J., Fried, E.I., Goodwin, G.M., 2020. Mood Homeostasis Before and During the Coronavirus Disease 2019 (COVID-19) Lockdown Among Students in the Netherlands. JAMA Psychiatry. https://doi.org/10.1001/jamapsychiatry.2020.2389

Twenge, J.M., Joiner, T.E., Rogers, M.L., Martin, G.N., 2017. Increases in Depressive Symptoms, Suicide-Related Outcomes, and Suicide Rates Among U.S. Adolescents After 2010 and Links to Increased New Media Screen Time: Clin. Psychol. Sci. https://doi.org/10.1177/2167702617723376

Verger, P., Combes, J.B., Kovess-Masfety, V., Choquet, M., Guagliardo, V., Rouillon, F., Peretti-Wattel, P., 2009. Psychological distress in first year university students: Socioeconomic and academic stressors, mastery and social support in young men and women. Soc. Psychiatry Psychiatr. Epidemiol. https://doi.org/10.1007/s00127-008-0486-y

Verger, P., Guagliardo, V., Gilbert, F., Rouillon, F., Kovess-Masfety, V., 2010. Psychiatric disorders in students in six French universities: 12-month prevalence, comorbidity, impairment and help-seeking. Soc. Psychiatry Psychiatr. Epidemiol. https://doi.org/10.1007/s00127-009-0055-z

Wang, C., Pan, R., Wan, X., Tan, Y., Xu, L., Ho, C.S., Ho, R.C., 2020. Immediate Psychological Responses and Associated Factors during the Initial Stage of the 2019 Coronavirus Disease (COVID-19) Epidemic among the General Population in China. Int. J. Environ. Res. Public. Health 17, 1729. https://doi.org/10.3390/ijerph17051729

WHO Coronavirus Disease (COVID-19) Dashboard | WHO Coronavirus Disease (COVID-19) Dashboard [WWW Document], n.d. URL https://covid19.who.int/table?tableChartType=heat (xaccessed 9.3.20).

