## Supplementary material for "Higher risk of mental health deterioration during the Covid-19 lockdown among students rather than non-students. The French Confins study"

**Table S1.** Additional characteristics of the study population, Confins cohort, France, 2020

| | Total N=2260 | | Students N=1335 | | Non-students N=925 | |
|---|---|---|---|---|---|---|
| **Depressive symptoms** | | | | | | |
| PHQ-9 score ≥15, n (%) | 272 | (12.0) | 204 | (15.3) | 68 | (7.4) |
| Minimal, n (%) | 1590 | (70.4) | 844 | (63.2) | 746 | (80.7) |
| Mild, n (%) | 362 | (16.0) | 264 | (19.8) | 98 | (10.6) |
| Moderate, n (%) | 194 | (8.6) | 143 | (10.7) | 51 | (5.5) |
| Severe, n (%) | 114 | (5.0) | 84 | (6.3) | 30 | (3.2) |
| **Anxiety symptoms** | | | | | | |
| GAD-7 score ≥ 15, n (%) | 172 | (7.6) | 126 | (9.4) | 46 | (5.0) |
| Minimal, n (%) | 1209 | (53.5) | 634 | (47.5) | 575 | (62.2) |
| Mild, n (%) | 531 | (23.5) | 336 | (25.2) | 195 | (21.1) |
| Moderate, n (%) | 300 | (13.3) | 203 | (15.2) | 97 | (10.5) |
| Severe, n (%) | 220 | (9.7) | 162 | (12.1) | 58 | (6.3) |
| **Perceived stress** | | | | | | |
| Perceived stress score ≥ 5, n (%) | 1086 | (56.6) | 736 | (61.8) | 350 | (48.1) |
| **Having a paid activity** (student job), n (%) (N=1100) | - | - | 219 | (19.9) | - | - |
| **Difficult financial situation during childhood**, n (%) (N=1100) | - | - | 420 | (38.2) | - | - |
| **Having a high professional position**, n (%) | - | - | - | - | 471 | (51.0) |

| | | | | | | |
|---|---|---|---|---|---|---|
| **Having a stable professional situation**, n (%) | - | - | - | - | 553 | (59.9) |
| **Working remotely during lockdown**, n (%) (N=924) | - | - | - | - | 429 | (46.4) |

**Table S2.** Sensitivity analysis for the effect of student status on mental health conditions during Covid-19 lockdown among young, estimated with multivariate logistic regression analysis, Confins cohort, France, 2020.

| | n | **Model 1** | | | **Model 2** | | | **Model 3** | | |
|---|---|---|---|---|---|---|---|---|---|---|
| | | **OR** | **IC 95%** | **p** | **OR** | **IC 95%** | **p** | **OR** | **IC 95%** | **p** |
| **Among imputed sample** | | | | | | | | | | |
| Depressive symptoms (PHQ-9 score ≥ 15) | 2260 | 1.30 | (0.90 ; 1.88) | 0.1568 | 1.41 | (0.95 ; 2.09) | 0.0852 | 1.35 | (0.90 ; 2.03) | 0.1444 |
| Full spectrum of depressive symptoms | 2260 | | | **0.0284** | | | **0.0269** | | | **0.0314** |
| Mild vs minimal | | **1.56** | **(1.12 ; 2.16)** | **0.0077** | **1.55** | **(1.11 ; 2.17)** | **0.0096** | **1.57** | **(1.11 ; 2.22)** | **0.0104** |
| Moderate vs minimal | | 1.33 | (0.87 ; 2.03) | 0.1887 | 1.42 | (0.91 ; 2.22) | 0.1232 | 1.32 | (0.83 ; 2.10) | 0.2340 |
| Severe vs minimal | | 1.45 | (0.84 ; 2.49) | 0.1827 | 1.62 | (0.90 ; 2.92) | 0.1066 | 1.69 | (0.92 ; 3.11) | 0.0896 |
| Anxiety symptoms (GAD-7 score ≥ 15) | 2260 | 1.37 | (0.88 ; 2.14) | 0.1650 | 1.44 | (0.90 ; 2.31) | 0.1297 | 1.53 | (0.94 ; 2.49) | 0.0902 |
| Full spectrum of depressive symptoms | 2260 | | | **0.0279** | | | **0.0238** | | | 0.0543 |
| Mild vs minimal | | 1.16 | (0.88 ; 1.53) | 0.2863 | 1.14 | (0.86 ; 1.51) | 0.3499 | 1.13 | (0.85 ; 1.51) | 0.4052 |
| Moderate vs minimal | | **1.46** | **(1.03; 2.08)** | **0.0338** | **1.49** | **(1.04 ; 2.15)** | **0.0313** | 1.38 | (0.95 ; 2.00) | 0.0940 |
| Severe vs minimal | | **1.68** | **(1.11 ; 2.55)** | **0.0143** | **1.82** | **(1.17 ; 2.84)** | **0.0078** | **1.81** | **(1.14 ; 2.86)** | **0.0114** |

| | | | | | | | | | |
|---|---|---|---|---|---|---|---|---|---|
| Perceived stress (score ≥ 5) | 1919 | **1.48** | **(1.16 ; 1.90)** | **0.0017** | **1.52** | **(1.18 ; 1.96)** | **0.0012** | **1.48** | **(1.14 ; 1.92)** | **0.0032** |
| **Among complete case sample** | | | | | | | | | | |
| Depressive symptoms (PHQ-9 score ≥ 10) | 1907 | **1.37** | **(1.03 ; 1.84)** | **0.0337** | **1.62** | **(1.17 ; 2.25)** | **0.0038** | **1.58** | **(1.13 ; 2.22)** | **0.0082** |
| Anxious symptoms (GAD-7 score ≥ 10) | 1907 | 1.36 | (0.99 ; 1.86) | 0.0543 | **1.53** | **(1.08 ; 2.16)** | **0.0173** | **1.46** | **(1.02 ; 2.09)** | **0.0388** |
| Suicidal thoughts (Yes versus No) | 1907 | 1.47 | (0.96 ; 2.27) | 0.0774 | **1.70** | **(1.04 ; 2.78)** | **0.0329** | 1.62 | (0.98 ; 2.68) | 0.0607 |
| Perceived stress (score ≥ 7) | 1907 | **1.61** | **(1.22 ; 2.13)** | **0.0009** | **1.71** | **(1.27 ; 2.31)** | **0.0004** | **1.68** | **(1.23 ; 2.29)** | **0.0010** |

Model 1 adjusted for sex and age; Model 2 adjusted for sex, age, being in a relationship, education level, working or studying for the medical sector, history of psychiatric disease, history of other disease at risk for Covid-19 ; Model 3 adjusted for sex, age, being in a relationship, education level, working or studying for the medical sector, history of psychiatric disease, history of other disease at risk for Covid-19, week of inclusion in the cohort, being in lockdown in a high-risk region, acquaintance or family with Covid-19 or Covid-19'suspicion, being in lockdown alone, lockdown accommodation with an outdoor space, having a pet, lockdown accommodation surface, physical activity during lockdown.

**Table S3.** Effect of college student status on mental health conditions during Covid-19 lockdown among young adults (≤ 30 years old), estimated with multivariate logistic regression analysis with imputed data on covariates, Confins cohort, France, 2020.

| | n | Model 1 | | | Model 2 | | | Model 3 | | |
|---|---|---|---|---|---|---|---|---|---|---|
| | | **OR** | **IC 95%** | **p** | **OR** | **IC 95%** | **p** | **OR** | **IC 95%** | **p** |
| Depressive symptoms (PHQ-9 score ≥ 10) | 1600 | 1.18 | (0.84 ; 1.68) | 0.3417 | **1.48** | **(1.01 ; 2.16)** | **0.0450** | **1.54** | **(1.03 ; 2.29)** | **0.0353** |
| Anxiety symptoms (GAD-7 score ≥ 10) | 1600 | 1.31 | (0.89 ; 1.91) | 0.1672 | **1.54** | **(1.02 ; 2.32)** | **0.0401** | **1.60** | **(1.04 ; 2.45)** | **0.0324** |
| Suicidal thoughts (yes vs no) | 1413 | 1.10 | (0.65 ; 1.87) | 0.7259 | 1.60 | (0.89 ; 2.90) | 0.1174 | 1.42 | (0.76 ; 2.63) | 0.2716 |
| Perceived stress (score ≥ 7) | 1413 | 1.34 | (0.95 ; 1.90) | 0.0989 | **1.61** | **(1.10 ; 2.35)** | **0.0136** | **1.59** | **(1.07 ; 2.34)** | **0.0205** |

Models adjusted for sex and age (18;20, 21;24 vs more than 25); Model 2 adjusted for sex, age (18;20, 21;24 vs more than 25), being in a relationship, education level, working or studying for the medical sector, history of psychiatric disease, history of other disease at risk for Covid-19 ; Model 3 adjusted for sex, age (18;20, 21;24 vs more than 25), being in a relationship, education level, working or studying for the medical sector, history of psychiatric disease, history of other disease at risk for Covid-19, week of inclusion in the cohort, being in lockdown in a high-risk region, acquaintance or family with Covid-19 or Covid-19'suspicion, being in lockdown alone, lockdown accommodation with an outdoor space, having a pet, lockdown accommodation surface, physical activity during lockdown.

**Table S4.** Factors associated with mental health conditions during Covid-19 lockdown, estimated with multivariate logistic regression analysis with imputed data on covariates, among students, Confins cohort, France, 2020.

| | **Depressive symptoms (n=1335)** | | | **Anxiety symptoms (n=1335)** | | | **Suicidal thoughts (n=1191)** | | | **Perceived stress (n=1191)** | | |
|---|---|---|---|---|---|---|---|---|---|---|---|---|
| | **OR** | **IC 95%** | **p** | **OR** | **IC 95%** | **p** | **OR** | **IC 95%** | **p** | **OR** | **IC 95%** | **p** |
| Female vs male | **1.74** | **(1.23 ; 2.45)** | **0.0016** | **1.88** | **(1.28 ; 2.76)** | **0.0014** | 1.24 | (0.72 ; 2.14) | 0.4433 | **1.65** | **(1.17 ; 2.34)** | **0.0045** |
| Age, in years | **0.92** | **(0.87 ; 0.96)** | **0.0007** | 0.97 | (0.93 ; 1.01) | 0.1727 | 0.97 | (0.91 ; 1.04) | 0.3556 | 0.97 | (0.93 ; 1.01) | 0.1917 |
| In a relationship vs no | 1.08 | (0.82 ; 1.41) | 0.5857 | **1.32** | **(0.99 ; 1.77)** | **0.0567** | 1.20 | (0.78 ; 1.85) | 0.3986 | 1.12 | (0.85 ; 1.47) | 0.4310 |
| Educational level | | | | | | | | | | | | |
| < Second-year university level vs > second-year university level | **2.04** | **(1.34 ; 3.11)** | **0.0009** | 1.45 | (0.93 ; 2.26) | 0.0971 | **3.00** | **(1.63 ; 5.53)** | **0.0004** | 1.46 | (0.95 ; 2.25) | 0.0804 |
| Second-year university level vs > second-year university level | **1.54** | **(1.06 ; 2.25)** | **0.0238** | **1.47** | **(0.99 ; 2.19)** | **0.0547** | 1.34 | (0.75 ; 2.40) | 0.3285 | **1.56** | **(1.07 ; 2.28)** | **0.0218** |
| Having a paid activity (student job) vs no | 0.86 | (0.60 ; 1.23) | 0.4027 | 1.13 | (0.77 ; 1.67) | 0.5263 | 0.87 | (0.50 ; 1.53) | 0.6363 | 1.16 | (0.82 ; 1.62) | 0.4005 |
| Studying for the medical sector vs no | **0.64** | **(0.48 ; 0.85)** | **0.0023** | **0.65** | **(0.48 ; 0.89)** | **0.0063** | 0.76 | (0.49 ; 1.19) | 0.2315 | 0.79 | (0.60 ; 1.04) | 0.0934 |
| History of psychiatric disease vs no | **2.80** | **(2.06 ; 3.82)** | **<0.0001** | **3.07** | **(2.23 ; 4.24)** | **<0.0001** | **5.08** | **(3.35 ; 7.71)** | **<0.0001** | **2.39** | **(1.76 ; 3.26)** | **<0.0001** |
| History of other disease at risk for Covid-19 vs no | 1.34 | (0.97 ; 1.85) | 0.0761 | 1.13 | (0.78 ; 1.62) | 0.5109 | 1.28 | (0.82 ; 1.99) | 0.2721 | 0.98 | (0.72 ; 1.32) | 0.8732 |

| | | | | | | | | | | | | |
|---|---|---|---|---|---|---|---|---|---|---|---|---|
| Difficult financial situation during childhood vs no | **1.45** | **(1.09 ; 1.92)** | **0.0108** | **1.55** | **(1.13 ; 2.12)** | **0.0071** | **1.78** | **(1.18 ; 2.70)** | **0.0063** | **1.51** | **(1.14 ; 2.01)** | **0.0044** |
| Week of inclusion in the cohort | | | | | | | | | | | | |
| W16-17 vs W14-15 | **1.60** | **(1.07 ; 2.39)** | **0.0214** | 1.36 | (0.88 ; 2.09) | 0.1710 | 1.01 | (0.52 ; 1.97) | 0.9653 | 1.33 | (0.83 ; 2.14) | 0.2346 |
| W18-19 vs W14-15 | **2.68** | **(1.68 ; 4.30)** | **<0.0001** | **2.44** | **(1.49 ; 4.02)** | **0.0004** | 1.07 | (0.51 ; 2.26) | 0.8581 | **1.89** | **(1.11 ; 3.22)** | **0.0187** |
| Being in lockdown in a high-risk region vs no | 0.92 | (0.60 ; 1.41) | 0.7083 | 0.77 | (0.47 ; 1.24) | 0.2833 | 0.86 | (0.43 ; 1.72) | 0.6698 | 0.88 | (0.57 ; 1.34) | 0.5437 |
| Acquaintance or family with Covid-19 or Covid-19 suspicion vs no | 1.17 | (0.90 ; 1.50) | 0.2403 | 1.17 | (0.89 ; 1.54) | 0.2616 | 0.99 | (0.66 ; 1.47) | 0.9455 | 1.07 | (0.83 ; 1.39) | 0.5919 |
| Being in lockdown alone vs no | **1.54** | **(1.04 ; 2.28)** | **0.0325** | 1.07 | (0.69 ; 1.64) | 0.7705 | 2.32 | (1.30 ; 4.16) | 0.0045 | 1.29 | (0.87 ; 1.91) | 0.2074 |
| Lockdown accommodation with an outdoor space vs no | 1.14 | (0.81 ; 1.61) | 0.4602 | 1.05 | (0.72 ; 1.51) | 0.8130 | **1.62** | **(0.96 ; 2.72)** | **0.0707** | 1.07 | (0.76 ; 1.52) | 0.6945 |
| Having a pet vs no | 1.02 | (0.78 ; 1.34) | 0.8918 | 1.12 | (0.84 ; 1.49) | 0.4448 | 1.31 | (0.85 ; 2.02) | 0.2225 | 1.08 | (0.82 ; 1.42) | 0.5903 |
| Lockdown accommodation surface (for 10 $m^2$) | **0.98** | **(0.97 ; 0.99)** | **0.0064** | 1.00 | (0.99 ; 1.00) | 0.3021 | **0.93** | **(0.89 ; 0.97)** | **0.0015** | 0.99 | (0.98 ; 1.00) | 0.0951 |
| Physical exercise during lockdown vs no | **0.61** | **(0.44 ; 0.83)** | **0.0019** | **0.58** | **(0.41 ; 0.80)** | **0.0012** | **0.54** | **(0.34 ; 0.84)** | **0.0068** | **0.61** | **(0.44 ; 0.84)** | **0.0022** |
| Being in lockdown at home vs no | **0.75** | **(0.57 ; 0.99)** | **0.0417** | 0.85 | (0.63 ; 1.14) | 0.2782 | 0.67 | (0.43 ; 1.06) | 0.0864 | 0.96 | (0.72 ; 1.28) | 0.7886 |

p: p-value for the Wald test or type 3 test for categorical variables.

**Table S5.** Factors associated with mental health conditions during Covid-19 lockdown, estimated with multivariate logistic regression analysis with imputed data on covariates, among non-students, Confins cohort, France, 2020.

| | **Depressive symptoms (n=925)** | | | **Anxiety symptoms (n=925)** | | | **Suicidal thoughts (n=728)** | | | **Perceived stress (n=728)** | | |
|---|---|---|---|---|---|---|---|---|---|---|---|---|
| | **OR** | **IC 95%** | **p** | **OR** | **IC 95%** | **p** | **OR** | **IC 95%** | **p** | **OR** | **IC 95%** | **p** |
| Female vs male | 1.14 | (0.70 ; 1.87) | 0.5902 | 0.98 | (0.61 ; 1.58) | 0.9495 | 0.75 | (0.37 ; 1.52) | 0.4228 | 1.24 | (0.79 ; 1.96) | 0.3518 |
| Age | **0.98** | **(0.96 ; 0.99)** | **0.0103** | 0.99 | (0.98 ; 1.01) | 0.5008 | 0.98 | (0.96 ; 1.01) | 0.2189 | 1.00 | (0.98 ; 1.01) | 0.5964 |
| In a relationship vs no | 0.67 | (0.40 ; 1.10) | 0.1144 | 0.60 | (0.36 ; 1.00) | 0.0519 | 0.95 | (0.43 ; 2.13) | 0.9077 | 0.71 | (0.43 ; 1.17) | 0.1843 |
| Educational level | | | | | | | | | | | | |
| < Second-year university level vs > second-year university level | **1.89** | **(1.05 ; 3.39)** | **0.0335** | 1.69 | (0.94 ; 3.04) | 0.0774 | 1.38 | (0.60 ; 3.20) | 0.4524 | 1.69 | (0.96 ; 2.97) | 0.0714 |
| Second-year university level vs > second-year university level | 0.94 | (0.53 ; 1.66) | 0.8349 | 0.97 | (0.54 ; 1.72) | 0.9128 | 0.42 | (0.15 ; 1.20) | 0.1046 | 1.04 | (0.60 ; 1.78) | 0.8996 |
| Having a high professional position vs no | **0.55** | **(0.33 ; 0.91)** | **0.0201** | 0.76 | (0.46 ; 1.26) | 0.2949 | 0.56 | (0.25 ; 1.26) | 0.1638 | 1.06 | (0.66 ; 1.71) | 0.7971 |
| Having a stable professional situation vs no | 0.69 | (0.45 ; 1.05) | 0.0817 | 0.77 | (0.50 ; 1.18) | 0.2294 | 0.99 | (0.51 ; 1.93) | 0.9848 | **0.61** | **(0.41 ; 0.91)** | **0.0166** |

| | | | | | | | | | | | | |
|---|---|---|---|---|---|---|---|---|---|---|---|---|
| Working for the medical sector vs no | 0.71 | (0.45 ; 1.13) | 0.1535 | 0.81 | (0.51 ; 1.28) | 0.3679 | 0.77 | (0.36 ; 1.64) | 0.4983 | 0.84 | (0.54 ; 1.29) | 0.4139 |
| History of psychiatric disease vs no | **3.10** | **(1.99 ; 4.81)** | **<0.0001** | **2.47** | **(1.56 ; 3.89)** | **0.0001** | **4.27** | **(2.28 ; 7.98)** | **<0.0001** | **2.25** | **(1.49 ; 3.39)** | **0.0001** |
| History of other disease at risk for Covid-19 vs no | 1.20 | (0.75 ; 1.94) | 0.4422 | 1.14 | (0.75 ; 1.75) | 0.5295 | 1.48 | (0.78 ; 2.80) | 0.2313 | 1.14 | (0.77 ; 1.69) | 0.5191 |
| Week of inclusion in the cohort | | | | | | | | | | | | |
| W16-17 vs W14-15 | 1.14 | (0.74 ; 1.75) | 0.5465 | 1.03 | (0.67 ; 1.59) | 0.8763 | 1.01 | (0.48 ; 2.14) | 0.9794 | 0.82 | (0.53 ; 1.26) | 0.3615 |
| W18-19 vs W14-15 | 1.10 | (0.51 ; 2.37) | 0.8085 | 1.18 | (0.57 ; 2.48) | 0.6527 | 2.60 | (0.98 ; 6.91) | 0.0548 | 1.35 | (0.70 ; 2.58) | 0.3715 |
| Being in lockdown in a high-risk region vs no | 1.51 | (0.95 ; 2.39) | 0.0800 | 1.14 | (0.72 ; 1.81) | 0.5778 | 0.81 | (0.37 ; 1.77) | 0.5912 | 1.21 | (0.78 ; 1.88) | 0.3905 |
| Acquaintance or family with Covid-19 or Covid-19 suspicion vs no | 1.31 | (0.88 ; 1.95) | 0.1888 | **1.58** | **(1.06 ; 2.37)** | **0.0258** | 0.54 | (0.28 ; 1.05) | 0.0679 | 1.43 | (0.98 ; 2.09) | 0.0661 |
| Being in lockdown alone vs no | 0.90 | (0.50 ; 1.63) | 0.7384 | 0.80 | (0.44 ; 1.45) | 0.4586 | 1.96 | (0.81 ; 4.74) | 0.1343 | 0.72 | (0.39 ; 1.31) | 0.2796 |
| Lockdown accommodation with an outdoor space vs no | 0.63 | (0.38 ; 1.04) | 0.0724 | 0.66 | (0.40 ; 1.10) | 0.1074 | 0.97 | (0.41 ; 2.28) | 0.9435 | 1.09 | (0.63 ; 1.88) | 0.7656 |
| Having a pet vs no | 1.06 | (0.70 ; 1.61) | 0.7805 | 0.94 | (0.62 ; 1.44) | 0.7835 | 0.90 | (0.47 ; 1.73) | 0.7609 | 1.17 | (0.79 ; 1.73) | 0.4346 |

| | | | | | | | | | | | | |
|---|---|---|---|---|---|---|---|---|---|---|---|---|
| Lockdown accommodation surface (for 10 $m^2$) | 0.98 | (0.95 ; 1.01) | 0.2236 | 0.99 | (0.96 ; 1.01) | 0.3023 | 0.99 | (0.97 ; 1.01) | 0.4282 | 1.00 | (0.99 ; 1.01) | 0.7207 |
| Physical exercise during lockdown vs no | **0.52** | **(0.32 ; 0.83)** | **0.0066** | **0.44** | **(0.27 ; 0.70)** | **0.0005** | 0.76 | (0.36 ; 1.58) | 0.4609 | **0.61** | **(0.38 ; 1.00)** | **0.0496** |
| Being in lockdown at home vs no | 1.88 | (0.85 ; 4.18) | 0.1200 | 1.29 | (0.62 ; 2.67) | 0.5008 | 0.61 | (0.24 ; 1.56) | 0.2990 | 1.45 | (0.77 ; 2.74) | 0.2522 |
| Working remotely during lockdown vs Non | 0.83 | (0.52 ; 1.34) | 0.4460 | 0.74 | (0.46 ; 1.19) | 0.2118 | 0.67 | (0.30 ; 1.47) | 0.3125 | 0.95 | (0.61 ; 1.49) | 0.8211 |

p: p-value for the Wald test or type 3 test for categorical variables.
